# Telehealth versus face-to-face delivery of speech language pathology services: a systematic review and meta-analysis

**DOI:** 10.1101/2024.04.08.24305455

**Authors:** Anna Mae Scott, Justin Clark, Magnolia Cardona, Tiffany Atkins, Ruwani Peiris, Hannah Greenwood, Rachel Wenke, Elizabeth Cardell, Paul Glasziou

## Abstract

**Background:** There is an increasing demand for the provision of speech language pathology (SLP) services via telehealth. Therefore, we systematically reviewed randomized controlled trials comparing telehealth to face-to-face provision of SLP services.

**Methods:** We searched Medline, Embase, and Cochrane, clinical trial registries, and conducted a citation analysis to identify trials. We included randomized trials comparing similar care delivered live via telehealth (phone or video), to face-to-face. Primary outcomes included: % syllables stuttered (%SS) (for individuals who stutter); change in sound pressure levels monologue (for individuals with Parkinson’s disease); and key function scores (for other areas). Where data were sufficient, mean differences were calculated.

**Results:** Nine randomized controlled trials were included; 8 evaluated video and 1 evaluated phone telehealth. Risk of bias was generally low or unclear, excepting blinding. There were no significant differences at any time-point up to 18 months for %SS (mean difference, MD 0.1, 95% CI –0.4 to 0.6, p=0.70). For people with Parkinson’s disease, there was no difference between groups in change in sound pressure levels (monologue) (MD 0.6, 95% CI –1.2 to 2.5, p=0.49). Four trials investigated interventions for speech sound disorder, voice disorder, and post-stroke dysphagia and aphasia; they found no differences between telehealth service delivery and face-to-face delivery.

**Conclusions:** Evidence suggests that the telehealth provision of SLP services may be a viable alternative to their provision face-to-face, particularly to people who stutter and people with Parkinson’s disease. The key limitation is the small number of randomized controlled trials, as well as evidence on the quality of life, well-being and satisfaction, and economic outcomes.

## BACKGROUND

Speech Language Pathologists play a vital role in the assessment and management of a range of communication disorders (e.g., speech, language voice, fluency) and swallowing disorders across the lifespan. These disorders can markedly affect individual’s quality of life, as they limit participation and social engagement,(1) resulting in an increased risk of depression and anxiety,(2) as well as developmental delays,(3) and higher risk of psychological morbidity in children.(2) However, when managed with ongoing therapy delivered by speech language pathologists, individuals with communication and swallowing disorders can achieve successful outcomes, and improved quality of life.

Due to concerns about geographic accessibility, or financial restrictions, some studies have explored the provision of speech language pathology services by means other than their traditional in-person delivery – including telehealth (also referred to as telecare or telemedicine). Telehealth involves the provision of healthcare services remotely, synchronously (live) or asynchronously, and using a broad range of information and communications technologies, such as video conferencing, teleconferencing, remote monitoring, mobile apps, and others.(4, 5)

The World Health Organization has promoted telehealth as a means of increasing accessibility, equity, quality and cost-effectiveness of health care services.(5) Patient acceptability of telehealth is often found to be high,(6) and patient satisfaction with consultations or treatment received via telehealth is often no different than with those received for face-to-face, across a range of conditions managed by allied healthcare providers, including PTSD,(7) depression,(8) anxiety,(9) management of musculoskeletal conditions(10) and others.(11) Telehealth delivery of speech language pathology in particular, has shown client acceptability and promising clinical outcomes, for example in vocal loudness and sentence intelligibility, (12) and in cost savings for health services.(13)

Evidence from systematic reviews of the effectiveness of telehealth in specific population subgroups is promising. For example, a systematic review evaluating the evidence for the effectiveness of voice therapy programs in adult populations, supported the use of telehealth as a service delivery model in speech language pathology for adults, although found that the evidence was limited in volume, most of the included studies lacked a control group, and meta-analyses were not able to be undertaken.(14) Another review, focusing on speech language pathology interventions delivered by telehealth to primary school-aged children with speech or language difficulties, included seven studies, showing similar improvements for children receiving interventions by telehealth and those receiving interventions in-person, similarly suggesting promising evidence in support of telehealth delivery in this group.(15) Finally, a review of telehealth assessment or interventions for individuals with autism spectrum disorder, found 14 United States-based studies utilizing a range of study designs, which suggested that telehealth delivery may be equivalent to those delivered face-to-face.(16)

As previous reviews focused on the provision of care by speech language pathologists to individuals with specific conditions or particular population subgroups, the volume and type of includable evidence was limited, and meta-analyses not possible. We therefore aimed to systematically review trials comparing the delivery of *any therapy* delivered by speech language pathologists via live telehealth, to its delivery face to face. We limited includable studies to randomized controlled trials only, to enable meta-analyses.

## METHODS

We aimed to find, appraise, and synthesize studies that compared any therapy delivered by a speech language pathologist via telehealth (video or telephone or both) to face to face consultations, for patients of all ages, and with any speech, language, fluency, voice, or swallowing disorder. This systematic review is reported following the Preferred Reporting Items for Systematic Reviews and Meta-Analyses (PRISMA) 2020 statement. The protocol for the systematic review was developed a priori, but it was not registered with PROSPERO.

### Inclusion criteria

#### Participants

We included studies of patients of all ages with persistent primary or secondary conditions seen by a speech language pathologist (including, but not limited to, speech, language, voice, fluency and swallowing disorders).

#### Intervention and comparator

We included studies of any interventions provided by speech language pathologists in primary care settings, in any country, by telehealth (telephone or video), comparing to their provision face-to-face. We included studies where the two groups received identical or nearly-identical therapy in terms of intensity, dose, frequency and content, by identical or nearly-identical health professionals. We excluded studies where services were provided asynchronously.

#### Outcomes (primary, secondary)

The primary outcomes and secondary outcomes were identified jointly with speech language pathologists, and differed by the condition addressed. For studies of individuals with stuttering, the outcomes included: % syllables stuttered (primary), and stuttering severity, time to complete treatment, satisfaction and quality of life measures (secondary outcomes). For studies of individuals with Parkinson’s disease, the primary outcome was the change in sound pressure levels monologue; and the secondary outcomes included: acoustic parameters, perceptual parameters, communication partner rating, satisfaction, and quality of life measures. For other studies, the outcomes included key function scores as reported in the study (primary); and time to complete treatment, satisfaction, and quality of life measures (secondary outcomes).

#### Study design

We included randomized controlled trials of any design (e.g. parallel, crossover, factorial, cluster). Systematic reviews were searched for any additional includable trials. We excluded all other study designs.

### Search strategies to identify studies

This review was conducted as part of a series of systematic reviews on the effectiveness of telehealth compared to face-to-face healthcare provision in primary care or allied care for a wide range of patient groups and conditions. Therefore, the search strings were deliberately very broad. We searched Medline (via PubMed), Embase (via Elsevier), and Cochrane (including CENTRAL), from inception until 22 June 2023. The complete search strings for all databases are provided in an Appendix.

On 18 October 2023, we conducted a forward (citing) and backwards (cited by) citation analysis using the SpiderCite tool.

No restrictions by language or publication date were imposed. We included study reports that were published in full; publications available as abstract only (e.g. as a conference abstract) were included if they had a clinical trial registry record, or other public report, with the additional information required for inclusion. Conference abstracts only with no additional information available were excluded.

### Study selection and screening

Pairs of authors (AMS, JC, MC, TA, RP, HG, PG) screened references independently, against the inclusion criteria – in title abstract and in full text. Any disagreements in decision about inclusion or exclusion were resolved by discussion or by adjudication by another author who was not a member of the original pair. Screening was conducted in either Endnote or Screenatron, as per each screener’s preference. The selection process was recorded in sufficient detail to complete a PRISMA flow diagram (see Figure 1).

**Figure 1:**
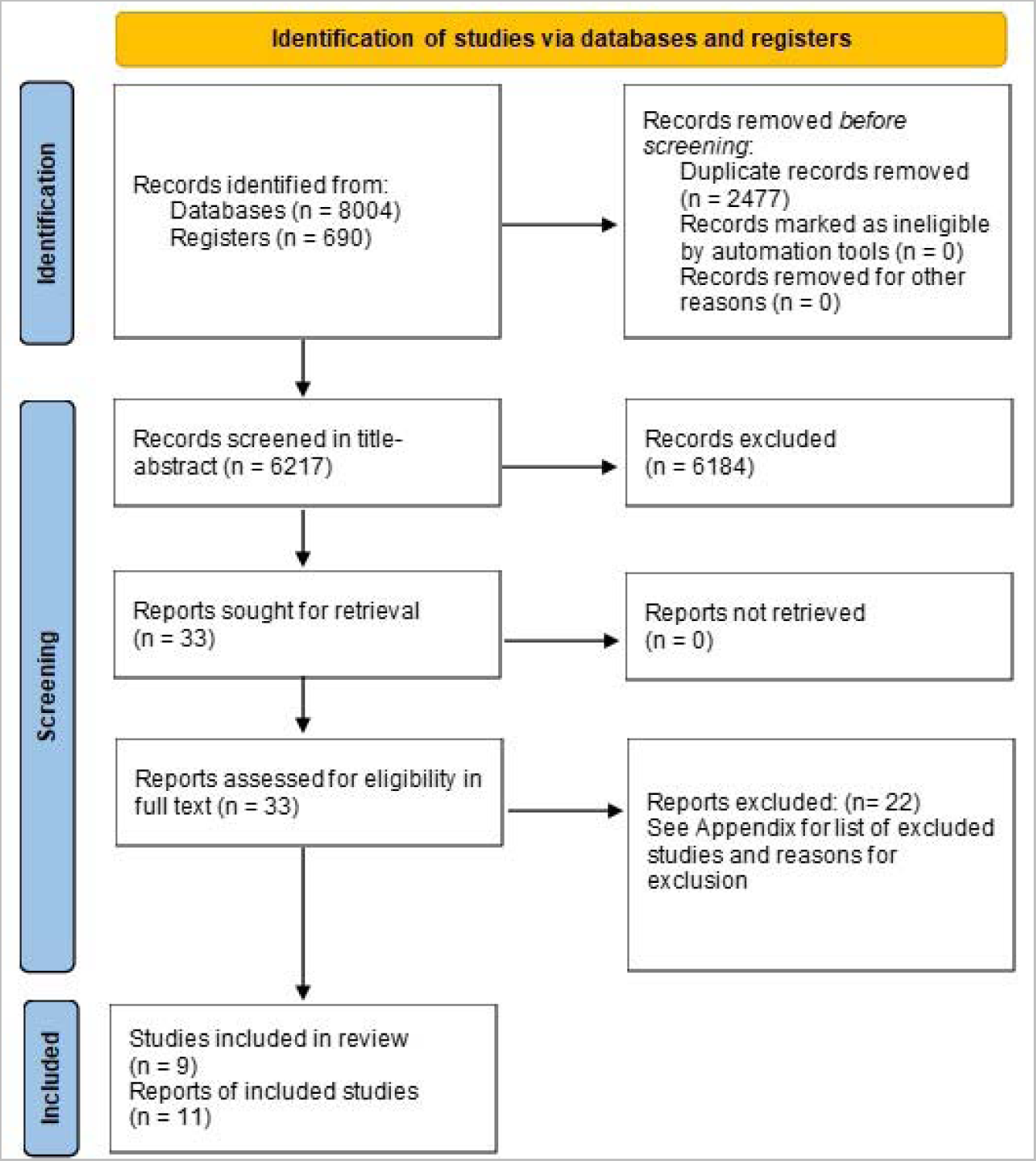
PRISMA Flow Diagram

**Figure 2:**
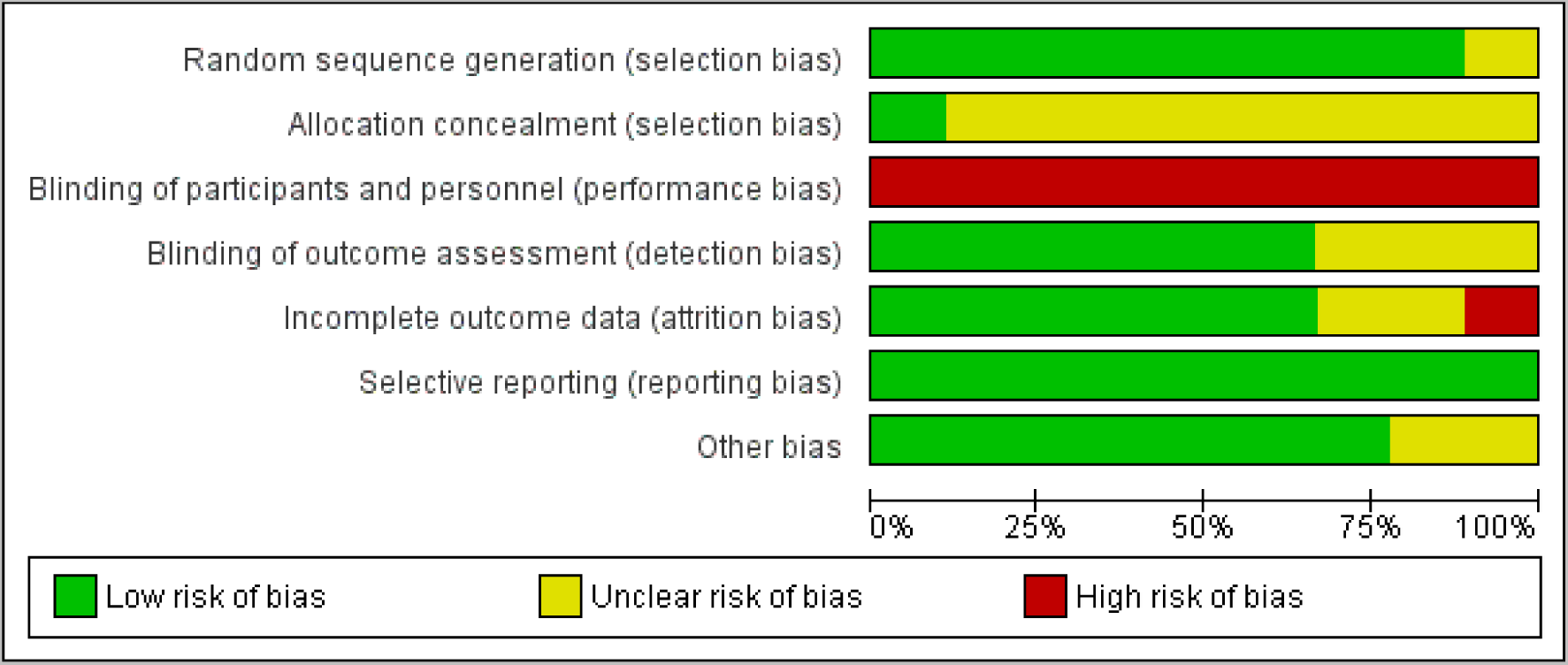
Risk of bias graph: review authors’ judgments about each risk of bias item presented as percentages across all included studies.

### Data extraction

We used a data extraction form to extract study data, which was piloted on 2 studies included in this review. From each study, we extracted information on: study characteristics (methods, participants, interventions, comparators, and outcomes), outcomes (primary and secondary) and data to inform the risk of bias judgments. Data were extracted by 4 authors independently (AMS, JC, MC, TA). Discrepancies were resolved by consensus, or reference to third author if required.

### Assessment of risk of bias in included studies

Paired review authors (AMS, JC, MC, TA) independently assessed the risk of bias for each included study using the Risk of Bias Tool 1, as outlined on the *Cochrane Handbook.*(17) Risk of Bias Tool 1 was used in preference to Risk of Bias Tool 2, as the former enables the evaluation of potential biases due to funding or conflict of interest, under “other bias” (domain 7).

Each potential source of bias was graded as low, high or unclear, supported by a quote from the relevant trial. The following domains were assessed:

1. Random sequence generation
2. Allocation concealment
3. Blinding of participants and personnel
4. Blinding of outcome assessment
5. Incomplete outcome data
6. Selective outcome reporting
7. Other bias (focusing on biases due to funding or conflict of interest).

All disagreements were resolved by discussion or by referring to a third author if required.

### Measurement of effect and data synthesis

*Review Manger 5.4* was used to calculate the treatment effect. For continuous outcomes (e.g. severity scores, satisfaction scores), we used mean difference for outcomes measured using the same scale across studies, or standardized mean difference, where outcomes were measured using different scales across studies. We undertook meta-analyses only when meaningful (i.e., when ≥2 studies or comparisons reported the same outcome).

Anticipating considerable heterogeneity, we used a random effects model. We used the I statistic to measure heterogeneity among the included trials. (17) Because we included fewer than 10 trials, we did not create a funnel plot to assess the publication bias.

The individual was used as the unit of analysis, where possible. However, where data on the number of individuals with outcomes of interest was not available, we extracted the information as it was presented (e.g., mean difference in scores). We did not contact investigators or study sponsors to provide missing data.

### Subgroup and sensitivity analyses

We conducted a subgroup analysis by time-point at which the outcome was reported, but not by study age groups or condition being treated due to few included studies. We had planned to conduct a sensitivity analysis by including versus excluding studies at high risk of bias, however, due to a low number of included studies and similarity of bias profiles, we were unable to do so.

## RESULTS

### Search results

Searches yielded in total 8694 references, resulting in 6217 references to screen in title-abstract after deduplication. We excluded 6184 references at the title-abstract screen, including 33 references for full text screen. 22 references were excluded (reasons for exclusion are provided in the Appendix), and 11 references corresponding to 9 trials were included in the review. (**Error! Reference source not found.**).

### Characteristics of included studies

We included 9 trials (11 references) comparing the provision of speech language pathology services via telehealth to face-to-face.(12, 18–26)(27) All were parallel, randomized controlled trials. Four trials took place in Australia, 3 trials in the United States, 1 in Taiwan, and 1 in Canada. Two trials evaluated the provision of speech pathology care for individuals with stuttering, 3 for patients with Parkinson’s disease, 2 for patients who were post-stroke, and 2 for patients with other conditions. Trials ranged in size from 14-69 participants, and followed patients from 1 week to 18 months, although four trials measured the outcomes at the completion of the trial, with no follow-up. The majority of trials compared the provision of services via video to face-to-face; 1 trial compared phone to face-to-face services. (**Error! Reference source not found.**)

### Risk of bias

Risk of bias for the included studies was generally low or unclear. Risk of bias was mostly low for random sequence generation. Most studies were rated at unclear risk of bias from allocation concealment, due to non-reporting whether concealment was used. All trials were at high risk of bias from blinding of participants and personnel, due to the nature of the compared interventions (telephone or video, versus face-to-face). Blinding of outcome assessment was mostly low or unclear – as was incomplete outcome data (although one trial was assessed at high risk). All trials were assessed at low risk of bias for selective reporting, and most were rated at low risk of bias due to other biases (assessed for potential biases due to conflicts of interest or funding sources). (**Error! Reference source not found.**)

### Telehealth versus face-to-face SLP for patients with stuttering

Two trials(18–20) reported on the provision of speech language pathology services for patients with stuttering.

### Percent syllables stuttered

For the primary outcome, percent syllables stuttered, there were no significant differences between the telehealth and the face-to-face group, immediately post-interventions (mean difference, MD – 0.17, 95% CI –2.18 to 1.84), at 6-9 months post-intervention (MD 0.65, 95% CI –0.21 to 1.51), or at 18 months post-intervention (MD 0.10, 95% CI –0.39 to 0.58). Heterogeneity was very low (I =0%). (Figure 3)

**Figure 3:**
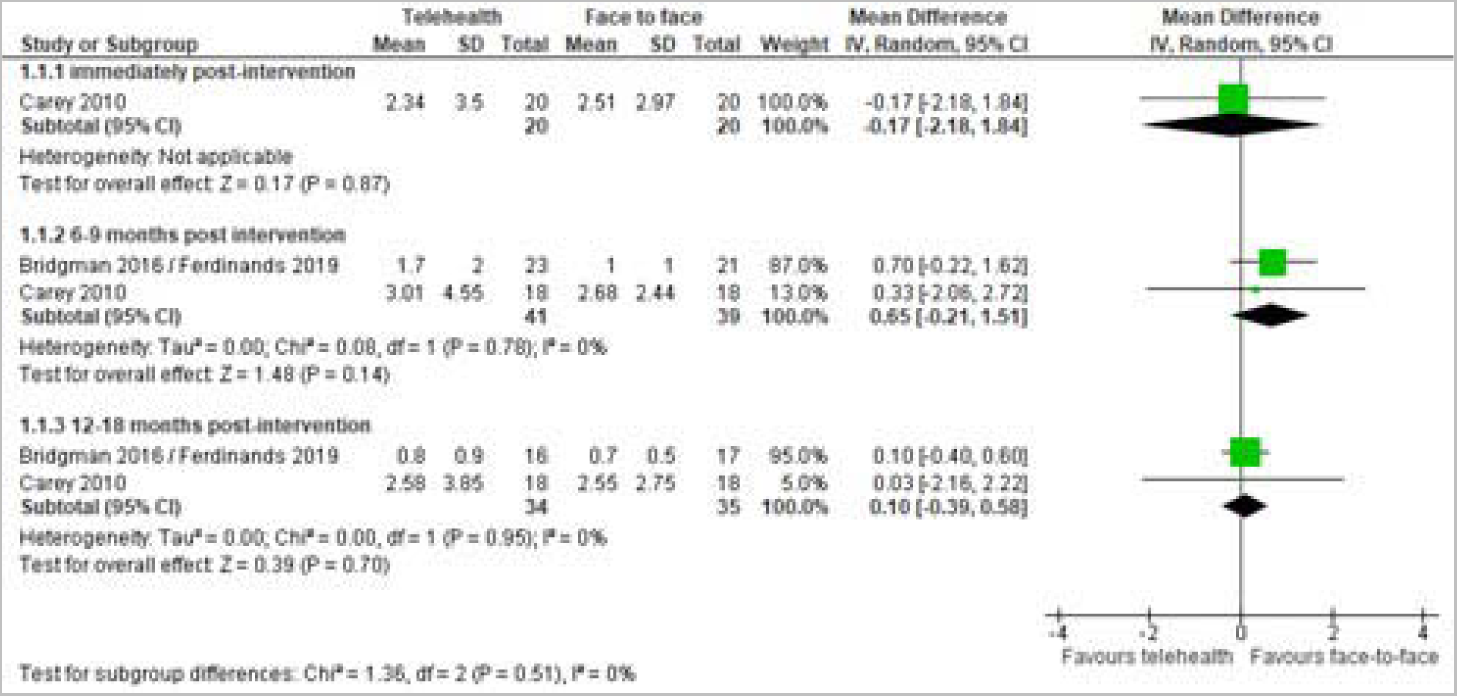
Telehealth vs. face-to-face for patients with stuttering: % syllables stuttered outcome

### Stuttering severity

For stuttering severity, 2 trials showed no differences between the telehealth and face-to-face groups. One trial with very young children (18, 20) reported no significant differences between the telehealth and face-to-face group either at 9 months post-intervention (MD: 0.1, 95% CI –0.56 to 0.49, P=0.88) or at 18 months post intervention (MD 0.1 95% CI –0.33 to 0.21, P=0.64). The trial with adults (19), assessing the self-reported stuttering severity on a scale of 1 to 9 (none to extremely severe, respectively), also found no difference, with the mean value of 2.3 in the telehealth group and 2.4 in the face-to-face group (p=0.7).

### Time to complete treatment

Both trials reported on the time required to complete treatment. One trial found that completion of stage 1 of the Lidcombe Program required a median of 25 weeks in both the telehealth and face-to-face group. However, there was a significance difference between groups in the mean duration (minutes) of consultations: face-to-face 40.4 min (standard deviation, SD 5.2) and telehealth 33.4 min (SD 4.7), p<0.001 (18, 20). Another trial found no difference in the mean number of speech pathologist contact hours to complete treatment (telehealth 617 minutes vs. face-to-face 774 minutes, p=0.17).(19)

### Satisfaction with treatment and/or outcomes

One trial with very young children (18, 20) measured parent satisfaction with their child’s fluency. There was no significant difference between the two groups either at 9 months post-intervention (p=0.54) or at 18 months post-intervention (p=0.74). Another trial with adults assessed treatment satisfaction, finding no difference between groups in the number of participants who described talking on the phone as ‘extremely easy’ (p=0.4); however, the telehealth group was significantly more frequently described as ‘extremely convenient’ (p=0.018). (19)

### Quality of life measures

Neither trial reported on quality of life measures.

### Telehealth versus face-to-face SLP for patients with Parkinson’s disease

Three trials(12, 21–23) reported on the provision of speech language pathology services for patients with Parkinson’s disease. All three compared the provision of Lee Silverman Voice Treatment via telehealth to its provision face to face.

### Change in sound pressure levels (monologue)

Three trials reported on the primary outcome, change in sound pressure level (monologue); two were meta-analyzable (12, 22, 23), showing no difference between the telehealth and face-to-face groups for change in sound pressure level (MD 0.64, 95% CI –1.20 to 2.48, p=0.49) (see Figure 4). Another trial (21) found no difference between the two groups (telehealth mean 67.91, face-to-face mean 69.5, p<0.17).

**Figure 4:**
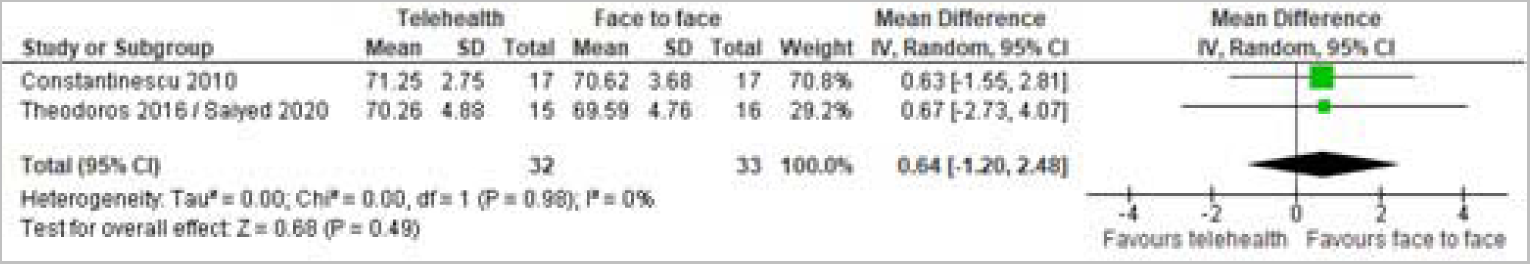
Telehealth vs. face-to-face for patients with Parkinson’s disease: change in sound pressure levels (monologue)

### Acoustic parameters

Two trials reported on sustained vowel phonation (SPL) (12, 22, 23), finding no difference between groups (MD –1.83, 95% CI –5.28 to 1.63, p=0.30). (Figure 5).

**Figure 5:**
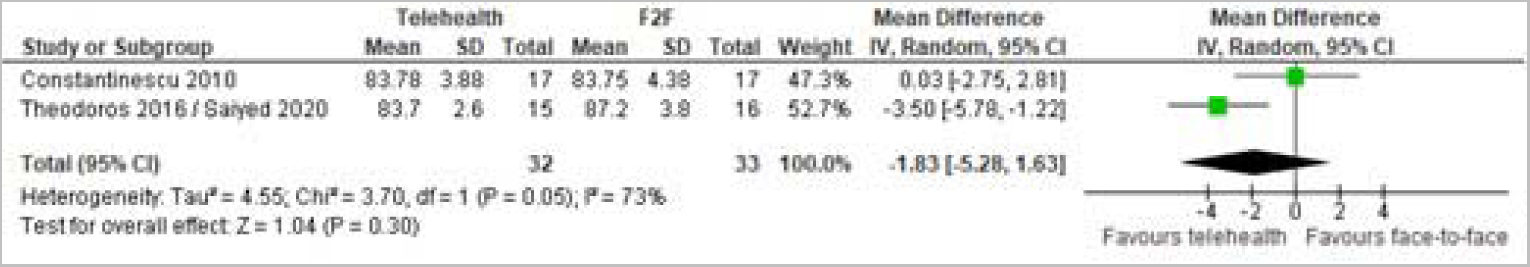
Telehealth vs. face-to-face for patients with Parkinson’s disease: sustained vowel phonation SPL

Three trials reported on the reading SPL outcome; two were meta-analyzable (12, 22, 23), and showed no difference between groups (MD –0.33, 95% CI –3.54 to 2.88, p=0.84) (Figure 6). One trial (21) found a significant difference between groups on the reading passage outcome (telehealth mean 70.05 dB, face-to-face mean 72.82 dB, p<0.05).

**Figure 6:**
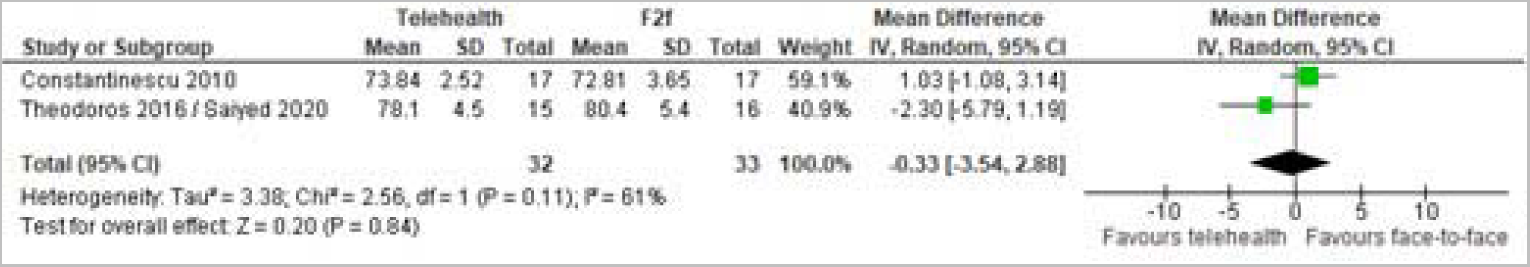
Telehealth vs. face-to-face for patients with Parkinson’s disease: reading SPL

Pooling two trials (12, 22, 23) that reported the difference in maximum fundamental frequency range showed no difference between the telehealth and face-to-face groups (SMD –0.12, 95% CI – 1.05 to 0.81, p=0.80). (Figure 7)

**Figure 7:**
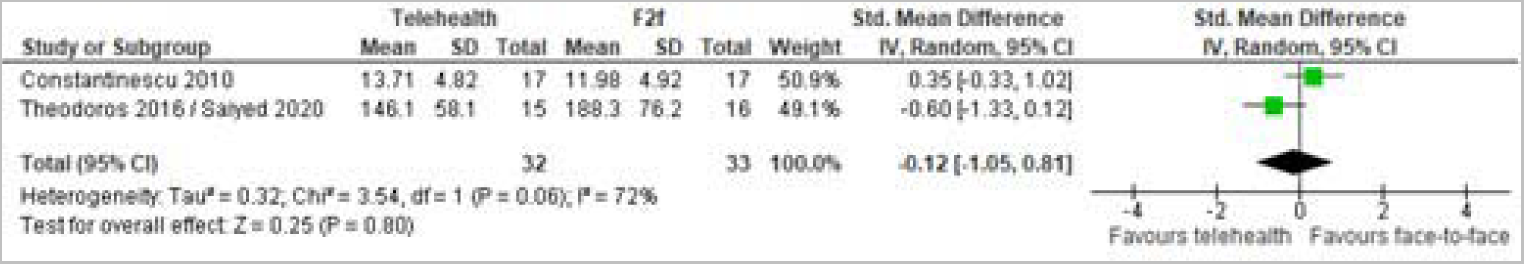
Telehealth vs. face-to-face for patients with Parkinson’s disease: maximum fundamental frequency change

### Perceptual parameters

Two trials reported on overall speech intelligibility (12, 22, 23), finding on difference between groups (MD –0.33, 95% CI –10.0 to 9.34, p=0.95) (Figure 8).

**Figure 8:**
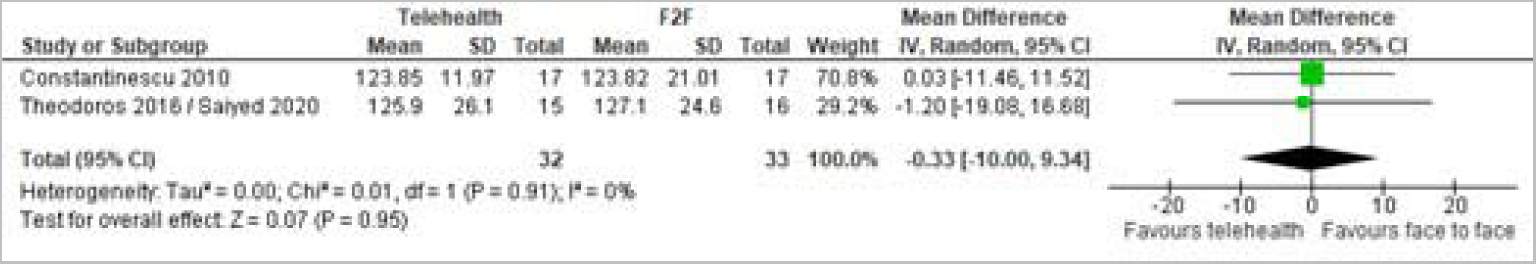
Telehealth vs. face-to-face for patients with Parkinson’s disease: overall speech intelligibility

Pooling two trials that reported on overall articulatory precision (12, 22, 23) showed no difference between telehealth and face-to-face groups (MD 4.28, 95% CI –16.39 to 24.95, p=0.69). (Figure 9)

**Figure 9:**
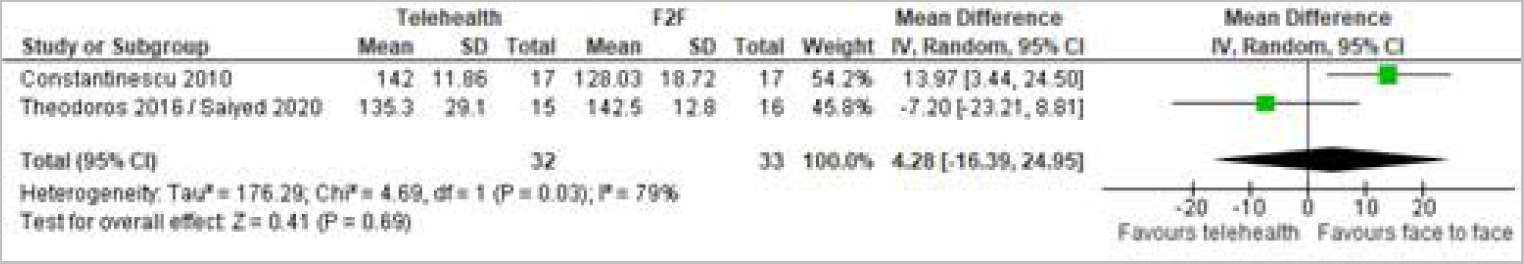
Telehealth vs. face-to-face for patients with Parkinson’s disease: overall articulatory precision

1 trial reported on percent sentence intelligibility. (12) The mean difference between the telehealth and face-to-face groups was not significant (MD 0.92, 95% CI –0.79 to 2.63, p=0.29).

Pooling two trials that reported on loudness showed no significant difference between telehealth and face-to-face groups (MD –4.13, 95% CI –16.39 to 8.12, p=0.51). (Figure 10)

**Figure 10:**
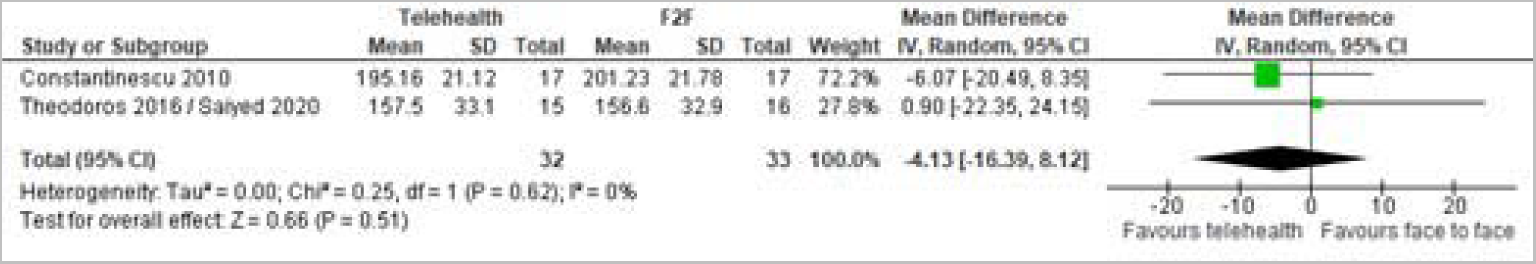
Telehealth vs. face-to-face for patients with Parkinson’s disease: loudness

### Communication partner rating

One trial (22, 23) reported on the communication partner rating, finding no difference between groups in the overall rating (MD –0.60, 95% CI –1.53 to 0.33, p=0.21).

### Quality of life/psychosocial measures

Quality of life and psychosocial measures were measured in one trial (22, 23).There was no significant difference between the telehealth and face-to-face group in the Dysarthria Impact Profile (DIP) total score (MD 4.90, 95% CI –10.38 to 20.18, p=0.53), and there was no significant difference between groups in the Parkinson’s Disease Questionnaire-39 Summary Index score (MD 0.30, 95% CI –7.92 to 8.52, p=0.94).

### Satisfaction

One trial (12) reported on the participant satisfaction in the telehealth (video) delivery of the Lee Silverman Voice Treatment, finding that the majority of participants were very happy (47.07%) or comfortable (47.07%) participating in the sessions, and the satisfaction with the video delivery of the treatment was high, with 18% of participants reporting being satisfied, 53% more than satisfied, and 29% very satisfied.

### Costs

One trial (22, 23) conducted an economic analysis of the cost of the 1-month speech language pathology program per patient. From the health system perspective, the costs for the telehealth program were slightly higher (mean $1076, standard deviation $71) than for the face-to-face program (mean $1020, SD $0). However, from the patient perspective, the costs were considerably lower for the telehealth program (mean $247, SD 99), than for the face-to-face program (mean $831, SD 570).

### Telehealth versus face-to-face SLP for patients with other conditions

Four other trials compared telehealth to face-to-face delivery of speech language pathology services: one trial for school children with speech sound impairments, (25) one trial for elderly persons with dysphonia,(26) one trial for patients with post-stroke dysphagia, (24) and one trial for patients with post-stroke aphasia.(27)

### School children with speech sound impairments

One trial (25) compared the delivery of speech sound intervention (personalized to each child’s needs) via video versus face-to-face.

Mean Goldman-Fristoe Test of Articulation 2 (GFTA-2) scores were not significantly different between the telehealth and face-to-face groups (MD –0.06, 95% CI –0.18 to 0.06, p=0.34). Mean listener judgments were also not significantly different between groups (p=0.057). The mean number of sessions attended was 9.3 in the telehealth group and 9.4 in the face-to-face group (p=0.73). Neither satisfaction nor quality of life outcomes were reported.

### Voice therapy for benign voice disorders in the elderly (dysphonia)

One trial (26) compared the delivery of voice therapy to the elderly with voice handicap index >10, via video to face-to-face.

There was no significant difference between groups post-treatment in Voice Handicap Index-10 scores, maximum phonation times (MPT), Jitter %, and smoothed cepstral peak prominence (CPP). There was, however, a significant difference in shimmer (p=0.04) and in noise-to-harmonic ratio (NHR) (p=0.01), both favoring the telehealth group (see Table 2).

**Table 1:** Characteristics of the included studies (n=9 Trials)

| Author<br>Year<br>Location | RCT<br>design | Follow up | Number of<br>participants<br>randomized | Participants | Age years<br>mean(SD) | Gender | Intervention | Telehealth: modality & dose | Comparator: modality & dose |
| --- | --- | --- | --- | --- | --- | --- | --- | --- | --- |
| ***Speech language pathology trials for stuttering*** |  |  |  |  |  |  |  |  |  |
| Bridgman 2016<br>&<br>Ferdinands<br>2019<br>Australia | Parallel<br>2-arm | 18 mo. | 49 (25 TH, 24<br>F2F) | Preschool<br>children who<br>stutter | NR (range 3-6) | NR | Lidcombe<br>Program | Video<br>45-60 min, 1x/week,<br>duration NR | F2F<br>45-60 min, 1x/week,<br>duration NR |
| Carey 2010<br>Australia | Parallel<br>2-arm | 12 mo. | 40 (20 TH, 20<br>F2F) | Adults who stutter | NR | Female: 7/40<br>(18%)<br>Male:<br>33/40 (82%) | Camperdown<br>Program | Phone<br>Varied, avg. 25 sessions,<br>30 weeks, 15.5 hrs total | F2F<br>Varied, avg 25<br>sessions, 30 weeks,<br>15.5 hrs total |
| ***Speech language pathology trials for patients with Parkinson's disease*** |  |  |  |  |  |  |  |  |  |
| Constantine-<br>scu 2010<br>Australia | Parallel<br>2-arm | N/A* | 34 (17 TH, 17<br>F2F) | Patients with<br>Parkinson's<br>disease | NR (range<br>54-85) | Female: 7/34<br>(21%)<br>Male:<br>27/34 (79%) | LSVT® | Video<br>1hr, 4x/week, 4 weeks | F2F<br>1hr, 4x/week, 4 weeks |
| Theodoros<br>2016<br>Sayied 2020<br>Australia | Parallel<br>2-arm** | N/A* | 31 (15 TH, 16<br>F2F) | Patients with<br>Parkinson's<br>disease | 71 (8.8) | Female: 10/31<br>(32%)<br>Male:<br>21/31 (68%) | LSVT® | Video<br>1hr, 4x/week, 4 weeks | F2F<br>1hr, 4x/week, 4 weeks |
| Covert 2018<br>USA | Parallel<br>2-arm | 1 week | 48 (18 TH, 18<br>F2F***) | Patients with<br>idiopathic<br>Parkinson's<br>disease | NR (range<br>54-87) | Female:***<br>2/36 (6%)<br>Male:.<br>34/36 (94%) | LSVT® | Video<br>1hr, 4x/week, 4 weeks | F2F<br>1hr, 4x/week, 4 weeks |
| ***Speech language pathology trials for patients post-stroke*** |  |  |  |  |  |  |  |  |  |
| Cassel 2017<br>USA | Parallel<br>2-arm | N/A* | 30 (15 TH, 15<br>F2F) | Patients with post-stroke dysphagia | NR (>65) | Female:<br>18/30 (60%)<br>Male:<br>12/30 (40%) | Dysphagia<br>therapy for safe<br>oral intake | Video<br>25-65 min, 1x only | F2F<br>25-65 min, 1x only |
| Meltzer 2018<br>Canada | Parallel 2-arm | 12 weeks | 44(22 TH, 22<br>F2F) | Patients with<br>post-stroke<br>aphasia | NR<br>Varied by<br>group; all<br>mean age<br>>60 | Female:<br>17/44 (39%)<br>Male:<br>27/44 (61%) | Communication<br>therapy | Video<br>1 hour<br>1x/week, 10 weeks | F2F<br>1 hour<br>1x/week, 10 weeks |
| ***Speech language pathology trials for other patient groups and/or conditions*** |  |  |  |  |  |  |  |  |  |
| Grogan-Johnson 2013<br>USA | Parallel<br>2-arm | N/A* | 14 (7 TH, 7<br>F2F) | School-aged<br>children with<br>speech sound<br>impairments | NR (range 6-10) | Female:<br>5/14 (36%)<br>Male:<br>9/14 (64%) | Speech sound<br>intervention<br>(personalized to<br>each child) | Video<br>30 min, 2x/week, 5 weeks | F2F<br>30 min,<br>2x/week, 5 weeks |
| Lin 2020<br>Taiwan | Parallel<br>2-arm | 8 weeks | 69 (33 TH, 36<br>F2F) | Elderly with voice<br>handicap index<br>>10 | NR (range<br>57-82) | Female:<br>33/69 (48%)<br>Male:<br>36/69 (52%) | Voice therapy | Video<br>30-45 min, 1x/week, 8<br>weeks | F2F<br>30-45 min, 1x/week, 8<br>weeks |

RCT=randomized controlled trial; mo.=month; TH=telehealth; F2F=face-to-face; LSVT: Lee Silverman Voice Treatment; NR=not reported; \*measures reported at the completion of the trial, no longer term follow-up; \*\*3<sup>rd</sup> arm (which was not randomized) is excluded from the present comparison; \*\*\* numbers randomized to each group unclear; 48 participants enrolled, 12 withdrew before finishing treatment; numbers reported are those analyzed for each group

**Table 2:** Telehealth vs. face-to-face for voice therapy for benign disorders in the elderly.

| Outcome | Telehealth |  |  | Face-to-face |  |  | MD [95% CI], p-value | Difference between groups |
| --- | --- | --- | --- | --- | --- | --- | --- | --- |
|  | mean | SD | N | mean | SD | N |  |  |
| VHI-10 | 16.8 | 8.94 | 25 | 13.46 | 9.95 | 24 | 3.34 [-1.96, 8.64], p=0.22 | Not significant |
| MPT | 9.83 | 3.2 | 25 | 10.15 | 5.85 | 24 | -0.32 [-2.98, 2.34], p=0.81 | Not significant |
| Jitter % | 1.73 | 1.16 | 25 | 2.63 | 2.01 | 24 | -0.90 [-1.82, 0.02], p=0.06 | Not significant |
| Shimmer | 0.44 | 0.17 | 25 | 0.67 | 0.53 | 24 | -0.23 [-0.45, -0.01], p=0.04 | Significant |
| NHR | 0.14 | 0.03 | 25 | 0.19 | 0.09 | 24 | -0.05 [-0.09, -0.01], p=0.01 | Significant |
| CPPs | 4.39 | 1.3 | 25 | 4.32 | 1.47 | 24 | 0.07 [-0.71, 0.85], p=0.86 | Not significant |

VHI-10: Voice Handicap Index-10; MPT: maximum phonation times; NHR: noise-to-harmonic ratio; CPPs: smoothed cepstral peak prominence

### Patients with post-stroke dysphagia

One trial (24) compared the delivery of instructional methods for dysphagia via video to face-to-face, to patients with post-stroke dysphagia. The trial found no significant difference between the telehealth and the face-to-face groups in swallowing ability, with 87% of participants in the telehealth group and 80% of the participants in the face-to-face groups achieving a goal of over 80% accuracy of 15 trials (difference not significant). There was also no difference between the telehealth and face-to-face groups in the mean scores of responses with cues (p=0.580) or without cues (p=0.870). The trial did not report nutritional measures (e.g. blood albumin), satisfaction with the mode of treatment delivery, or quality of life outcomes.

### Patients with post-stroke aphasia

One non-inferiority trial(27) compared communication therapy for patients with post-stroke aphasia via video versus face-to-face. There were no significant differences in reduction of aphasia severity as there was a 1.1 point average difference (90%CI: –2.05 to 4.26) in favor of telehealth compared to face-to-face. However, there was a significant difference between groups in favor of face-to-face (p=0.03) for levels of self-rated communication confidence as pre-to-post gains for telehealth (n=14, mean gain=2.18, SD=1.64) were not as high, compared to face-to-face (n=14, mean gain=4.79, SD=3.72). However, there were no statistically significant differences between groups in functional competence, as judged by the communication partner (p=0.83).

## DISCUSSION

This systematic review identified nine randomized clinical trials comparing the effectiveness of speech language pathology delivered by telehealth to conventional face-to-face delivery, for the management of persistent communication disorders in people with dysarthria following Parkinson’s disease, stuttering, and other conditions. Our findings indicate that speech language pathology services delivered via telehealth provide similar outcomes compared to services delivered face to face.

The results support favorable findings from earlier systematic reviews of telehealth in speech language pathology populations. (14, 28) Specifically, the present review confirmed that telehealth is no different to face-to-face approach for the provision of Lee Silverman Voice Treatment (LSVT®) in improving speech and wellbeing outcomes in Parkinson’s Disease, consistently with earlier efficacy studies of face-to-face therapy using the LSVT®.(29–31) As up to 90% of individuals with Parkinson’s Disease present with dysarthria (PD),(32) having accessible services via telehealth may have important implications for the communication and wellbeing of this population.

Similarly, meta-analyses of two trials confirmed similarity of telehealth vs. face-to-face treatment for young children who stutter. Systematic reviews targeting telehealth stuttering management by Lowe et al.(33) and McGill et al.(34) have similarly demonstrated positive client outcomes across the lifespan with evidence for the Lidcombe Program (young children), Camperdown program (adolescents, adults), and integrated approaches (young children, school-aged children, adults). Lowe et al(33) noted that telehealth delivery for very young children required more clinical hours than in-clinic face-to-face treatment, however, the reported Lidcombe Program RCT(18) did not show this. It must be noted that with the exception of the two trials reported in the current review(18, 19), all other studies have not had the rigor of being a RCT and most have had low numbers of participants (i.e., 1-6).

The present review is the first to include only randomized controlled trials comparing live telehealth (via video or phone) to face-to-face delivery for speech language pathology. It is a limitation in that this excluded a number of observational studies that have evaluated telehealth service delivery and show positive outcomes in populations frequently seen by speech language pathologists including aphasia, primary progressive aphasia and cognitive-communication disorders, were excluded from the review.(35–40) On the other hand, the restriction of includable studies is a strength in terms of allowing the meta-analyses to have been conducted. The study also excluded studies that provided asynchronous treatment, which excluded trials comparing face to face with asynchronous treatment modalities. Finally, we had not pre-specified the extraction of economic outcomes for the included studies, however, as one of the included studies (22, 23) conducted an economic analysis alongside the trial, we included those results.

Despite the currently limited volume of randomized controlled trial evidence which compares live telehealth to face-to-face provision of care by speech language pathologists, the emerging picture is that telehealth is a viable service delivery option. It merits acknowledging that currently, the randomized trial evidence for this comparison is limited to patients with stuttering, patients with Parkinson’s disease, and several others. Identified studies also included mostly male participants, with female participants ranging from 6% to 60% (median 32%), which limits generalizability of the findings. Nevertheless, from a healthcare provider perspective, telehealth models may in some cases increase attendance and adherence rates, and also enable speech pathology clients greater flexibility and more equitable access to services,(21) with comparable satisfaction rates to face-to-face services.(24)

## Data Availability

All data produced in the present work are contained in the manuscript

## Appendix 1 – Searches

**Searches run to 22/06/2023**

### RCT searches

#### PubMed

(“Telemedicine”[Mesh] OR “Videoconferencing”[Mesh] OR Telehealth[tiab] OR Telemedicine[tiab] OR Videoconferencing[tiab] OR ((Telephone[tiab]) AND (Consultation[tiab] OR face-to-face[tiab] OR in-person[tiab])) OR telephone-delivered[tiab]) AND

(“Primary Health Care”[Mesh] OR “General Practice”[Mesh] OR rehabilitation[sh] OR “Outpatients”[Mesh] OR “Speech Therapy”[Mesh] OR Outpatient[tiab] OR “Primary health”[tiab] OR “Primary care”[tiab] OR “General practice”[tiab] OR “General practices”[tiab] OR “General practitioners”[tiab] OR “General practitioner”[tiab] OR “Family practice”[tiab] OR Physician[tiab] OR Physicians[tiab] OR Clinician[tiab] OR Clinicians[tiab] OR Therapist[tiab] OR Nurse[tiab] OR Nurses[tiab] OR Physiotherapist[tiab] OR Rehabilitation[tiab] OR Diabetes[tiab] OR Diabetic[tiab] OR Asthma[tiab] OR Depression[tiab] OR “Ïrritable bowel”[tiab] OR IBS[tiab] OR PTSD[tiab] OR “Chronic fatigue”[tiab])

AND

((Face to face[tiab]) OR “Usual care”[tiab] OR Visits[tiab] OR Visit[tiab] OR In-person[tiab] OR “In person”[tiab] OR ((Clinic[tiab] OR Centre[tiab] OR Home[tiab]) AND (Based[tiab] OR Contact[tiab])) OR Conventional[tiab] OR “Practice-based”[tiab] OR “Practice based”[tiab] OR Traditional[tiab] OR “Standard care”[tiab] OR Homecare[tiab] OR ((Routine[tiab] OR Home[tiab]) AND (Care[tiab])))

AND

(“Delivery of Health Care”[Mesh] OR Delivery[tiab] OR Delivered[tiab] OR Via[tiab] OR Received[tiab])

AND

(“Treatment Outcome”[Mesh] OR “Patient Satisfaction”[Mesh] OR Therapy[sh] OR Diagnosis[sh] OR “Clinical outcomes”[tiab] OR Treatment[tiab] OR Diagnostic[tiab] OR Efficacy[tiab])

AND

(Randomized controlled trial[pt] OR controlled clinical trial[pt] OR randomized[tiab] OR randomised[tiab] OR placebo[tiab] OR “drug therapy”[sh] OR randomly[tiab] OR trial[tiab] OR groups[tiab])

NOT□

(Animals[Mesh] not (Animals[Mesh] and Humans[Mesh]))

NOT

(“Case Reports”[pt] OR Editorial[pt] OR Letter[pt] OR Meta-Analysis[pt] OR “Observational Study”[pt] OR “Systematic Review”[pt] OR “Case Report”[ti] OR “Case series”[ti] OR Meta-Analysis[ti] OR “Meta Analysis”[ti] OR “Systematic Review”[ti] OR “Systematic Literature Review”[ti] OR “Qualitative study”[ti] OR Protocol[ti])

#### CENTRAL

([mh Telemedicine] OR [mh Videoconferencing] OR Telehealth:ti,ab OR Telemedicine:ti,ab OR Videoconferencing:ti,ab OR ((Telephone:ti,ab) AND (Consultation:ti,ab OR “ face to face”:ti,ab OR “in person”:ti,ab)) OR “telephone delivered”:ti,ab)

AND

([mh “Primary Health Care”] OR [mh “General Practice”] OR [mh Outpatients] OR [mh “Speech Therapy”] OR Outpatient:ti,ab OR “Primary health”:ti,ab OR “Primary care”:ti,ab OR “General practice”:ti,ab OR “General practices”:ti,ab OR “General practitioners”:ti,ab OR “General practitioner”:ti,ab OR “Family practice”:ti,ab OR Physician:ti,ab OR Physicians:ti,ab OR Clinician:ti,ab OR Clinicians:ti,ab OR Therapist:ti,ab OR Nurse:ti,ab OR Nurses:ti,ab OR Physiotherapist:ti,ab OR Rehabilitation:ti,ab OR Diabetes:ti,ab OR Diabetic:ti,ab OR

Asthma:ti,ab OR Depression:ti,ab OR “Ïrritable bowel”:ti,ab OR IBS:ti,ab OR PTSD:ti,ab OR “Chronic fatigue”:ti,ab)

AND

((“Face to face”:ti,ab) OR “Usual care”:ti,ab OR Visits:ti,ab OR Visit:ti,ab OR “In person”:ti,ab OR ((Clinic:ti,ab OR Centre:ti,ab OR Home:ti,ab) AND (Based:ti,ab OR Contact:ti,ab)) OR Conventional:ti,ab OR “Practice based”:ti,ab OR Traditional:ti,ab OR “Standard care”:ti,ab OR Homecare:ti,ab OR ((Routine:ti,ab OR Home:ti,ab) AND (Care:ti,ab)))

AND

([mh “Delivery of Health Care”] OR Delivery:ti,ab OR Delivered:ti,ab OR Via:ti,ab OR Received:ti,ab)

AND

([mh “Treatment Outcome”] OR [mh “Patient Satisfaction”] OR “Clinical outcomes”:ti,ab OR Treatment:ti,ab OR Diagnostic:ti,ab OR Efficacy:ti,ab)

#### Embase

(‘Telemedicine’/exp OR ‘Videoconferencing’/exp OR Telehealth:ti,ab OR Telemedicine:ti,ab OR Videoconferencing:ti,ab OR ((Telephone:ti,ab) AND (Consultation:ti,ab OR face-to-face:ti,ab OR in-person:ti,ab)) OR telephone-delivered:ti,ab)

AND

(‘Primary Health Care’/exp OR ‘General Practice’/exp OR ‘Outpatient’/exp OR ‘Speech Therapy’/exp OR Outpatient:ti,ab OR “Primary health”:ti,ab OR “Primary care”:ti,ab OR “General practice”:ti,ab OR “General practices”:ti,ab OR “General practitioners”:ti,ab OR “General practitioner”:ti,ab OR “Family practice”:ti,ab OR Physician:ti,ab OR Physicians:ti,ab OR Clinician:ti,ab OR Clinicians:ti,ab OR Therapist:ti,ab OR Nurse:ti,ab OR Nurses:ti,ab OR Physiotherapist:ti,ab OR Rehabilitation:ti,ab OR Diabetes:ti,ab OR Diabetic:ti,ab OR Asthma:ti,ab OR Depression:ti,ab OR “Ïrritable bowel”:ti,ab OR IBS:ti,ab OR PTSD:ti,ab OR “Chronic fatigue”:ti,ab)

AND

((“Face to face”:ti,ab) OR “Usual care”:ti,ab OR Visits:ti,ab OR Visit:ti,ab OR In-person:ti,ab OR “In person”:ti,ab OR ((Clinic:ti,ab OR Centre:ti,ab OR Home:ti,ab) AND (Based:ti,ab OR Contact:ti,ab)) OR Conventional:ti,ab OR Practice-based:ti,ab OR “Practice based”:ti,ab OR Traditional:ti,ab OR “Standard care”:ti,ab OR Homecare:ti,ab OR ((Routine:ti,ab OR Home:ti,ab) AND (Care:ti,ab)))

AND

(‘health care delivery’/exp OR Delivery:ti,ab OR Delivered:ti,ab OR Via:ti,ab OR Received:ti,ab)

AND

(‘Treatment Outcome’/exp OR ‘Patient Satisfaction’/exp OR “Clinical outcomes”:ti,ab OR Treatment:ti,ab OR Diagnostic:ti,ab OR Efficacy:ti,ab)

AND

(random* OR factorial OR crossover OR placebo OR blind OR blinded OR assign OR assigned OR allocate OR allocated OR ‘crossover procedure’/exp OR ‘double-blind procedure’/exp OR ‘randomized controlled trial’/exp OR ‘single-blind procedure’/exp NOT (‘animal’/exp NOT (‘animal’/exp AND ‘human’/exp)))

□AND [embase]/lim

### Systematic Review searches

#### PubMed

(“Telemedicine”[Mesh] OR “Videoconferencing”[Mesh] OR Telehealth[tiab] OR Telemedicine[tiab] OR Videoconferencing[tiab] OR ((Telephone[tiab]) AND (Consultation[tiab] OR face-to-face[tiab] OR in-person[tiab])) OR telephone-delivered[tiab]) AND

(“Primary Health Care”[Mesh] OR “General Practice”[Mesh] OR rehabilitation[sh] OR “Outpatients”[Mesh] OR “Speech Therapy”[Mesh] OR Outpatient[tiab] OR “Primary health”[tiab] OR “Primary care”[tiab] OR “General practice”[tiab] OR “General practices”[tiab] OR “General practitioners”[tiab] OR “General practitioner”[tiab] OR “Family practice”[tiab] OR Physician[tiab] OR Physicians[tiab] OR Clinician[tiab] OR Clinicians[tiab] OR Therapist[tiab] OR Nurse[tiab] OR Nurses[tiab] OR Physiotherapist[tiab] OR Rehabilitation[tiab] OR Diabetes[tiab] OR Diabetic[tiab] OR Asthma[tiab] OR Depression[tiab] OR “Ïrritable bowel”[tiab] OR IBS[tiab] OR PTSD[tiab] OR “Chronic fatigue”[tiab])

AND

((Face to face[tiab]) OR “Usual care”[tiab] OR Visits[tiab] OR Visit[tiab] OR In-person[tiab] OR “In person”[tiab] OR ((Clinic[tiab] OR Centre[tiab] OR Home[tiab]) AND (Based[tiab] OR Contact[tiab])) OR Conventional[tiab] OR “Practice-based”[tiab] OR “Practice based”[tiab] OR Traditional[tiab] OR “Standard care”[tiab] OR Homecare[tiab] OR ((Routine[tiab] OR Home[tiab]) AND (Care[tiab])))

AND

(“Delivery of Health Care”[Mesh] OR Delivery[tiab] OR Delivered[tiab] OR Via[tiab] OR Received[tiab])

AND

(“Treatment Outcome”[Mesh] OR “Patient Satisfaction”[Mesh] OR Therapy[sh] OR Diagnosis[sh] OR “Clinical outcomes”[tiab] OR Treatment[tiab] OR Diagnostic[tiab] OR Efficacy[tiab])

AND

(Meta-Analysis[pt] OR “Systematic Review”[pt] OR Meta-Analysis[ti] OR “Meta Analysis”[ti] OR “Systematic Review”[ti] OR “Systematic Literature Review”[ti])

NOT

(“Case Reports”[pt] OR Editorial[pt] OR Letter[pt] OR “Observational Study”[pt] OR “Case Report”[ti] OR “Case series”[ti] OR “Qualitative study”[ti] OR Protocol[ti])

#### CDSR via the Cochrane Library

([mh Telemedicine] OR [mh Videoconferencing] OR Telehealth:ti,ab OR Telemedicine:ti,ab OR Videoconferencing:ti,ab OR ((Telephone:ti,ab) AND (Consultation:ti,ab OR “ face to face”:ti,ab OR “in person”:ti,ab)) OR “telephone delivered”:ti,ab)

AND

([mh “Primary Health Care”] OR [mh “General Practice”] OR [mh Outpatients] OR [mh “Speech Therapy”] OR Outpatient:ti,ab OR “Primary health”:ti,ab OR “Primary care”:ti,ab OR “General practice”:ti,ab OR “General practices”:ti,ab OR “General practitioners”:ti,ab OR “General practitioner”:ti,ab OR “Family practice”:ti,ab OR Physician:ti,ab OR Physicians:ti,ab OR Clinician:ti,ab OR Clinicians:ti,ab OR Therapist:ti,ab OR Nurse:ti,ab OR Nurses:ti,ab OR Physiotherapist:ti,ab OR Rehabilitation:ti,ab OR Diabetes:ti,ab OR Diabetic:ti,ab OR Asthma:ti,ab OR Depression:ti,ab OR “Ïrritable bowel”:ti,ab OR IBS:ti,ab OR PTSD:ti,ab OR “Chronic fatigue”:ti,ab)

AND

((“Face to face”:ti,ab) OR “Usual care”:ti,ab OR Visits:ti,ab OR Visit:ti,ab OR “In person”:ti,ab OR ((Clinic:ti,ab OR Centre:ti,ab OR Home:ti,ab) AND (Based:ti,ab OR Contact:ti,ab)) OR Conventional:ti,ab OR “Practice based”:ti,ab OR Traditional:ti,ab OR “Standard care”:ti,ab OR Homecare:ti,ab OR ((Routine:ti,ab OR Home:ti,ab) AND (Care:ti,ab)))

AND

([mh “Delivery of Health Care”] OR Delivery:ti,ab OR Delivered:ti,ab OR Via:ti,ab OR Received:ti,ab)

AND

([mh “Treatment Outcome”] OR [mh “Patient Satisfaction”] OR “Clinical outcomes”:ti,ab OR Treatment:ti,ab OR Diagnostic:ti,ab OR Efficacy:ti,ab)

#### Embase

(‘Telemedicine’/exp OR ‘Videoconferencing’/exp OR Telehealth:ti,ab OR Telemedicine:ti,ab OR Videoconferencing:ti,ab OR ((Telephone:ti,ab) AND (Consultation:ti,ab OR face-to-face:ti,ab OR in-person:ti,ab)) OR telephone-delivered:ti,ab)

AND

(‘Primary Health Care’/exp OR ‘General Practice’/exp OR ‘Outpatient’/exp OR ‘Speech Therapy’/exp OR Outpatient:ti,ab OR “Primary health”:ti,ab OR “Primary care”:ti,ab OR “General practice”:ti,ab OR “General practices”:ti,ab OR “General practitioners”:ti,ab OR “General practitioner”:ti,ab OR “Family practice”:ti,ab OR Physician:ti,ab OR Physicians:ti,ab OR Clinician:ti,ab OR Clinicians:ti,ab OR Therapist:ti,ab OR Nurse:ti,ab OR Nurses:ti,ab OR Physiotherapist:ti,ab OR Rehabilitation:ti,ab OR Diabetes:ti,ab OR Diabetic:ti,ab OR Asthma:ti,ab OR Depression:ti,ab OR “Ïrritable bowel”:ti,ab OR IBS:ti,ab OR PTSD:ti,ab OR “Chronic fatigue”:ti,ab)

AND

((“Face to face”:ti,ab) OR “Usual care”:ti,ab OR Visits:ti,ab OR Visit:ti,ab OR In-person:ti,ab OR “In person”:ti,ab OR ((Clinic:ti,ab OR Centre:ti,ab OR Home:ti,ab) AND (Based:ti,ab OR Contact:ti,ab)) OR Conventional:ti,ab OR Practice-based:ti,ab OR “Practice based”:ti,ab OR Traditional:ti,ab OR “Standard care”:ti,ab OR Homecare:ti,ab OR ((Routine:ti,ab OR Home:ti,ab) AND (Care:ti,ab)))

AND

(‘health care delivery’/exp OR Delivery:ti,ab OR Delivered:ti,ab OR Via:ti,ab OR Received:ti,ab)

AND

(‘Treatment Outcome’/exp OR ‘Patient Satisfaction’/exp OR “Clinical outcomes”:ti,ab OR Treatment:ti,ab OR Diagnostic:ti,ab OR Efficacy:ti,ab)

AND

([cochrane review]/lim OR [systematic review]/lim OR [meta analysis]/lim OR ((Search:ti,ab OR Searched:ti,ab) AND (PubMed:ti,ab OR MEDLINE:ti,ab)) OR (Systematic:ti,ab AND Review:ti,ab) OR ‘Meta analysis’:ti,ab OR Meta-analysis:ti,ab OR Review:ti OR ((Systematically:ti,ab OR Reviewed:ti,ab) AND (literature:ti,ab)))

## Appendix 2 – Table of Excluded Studies

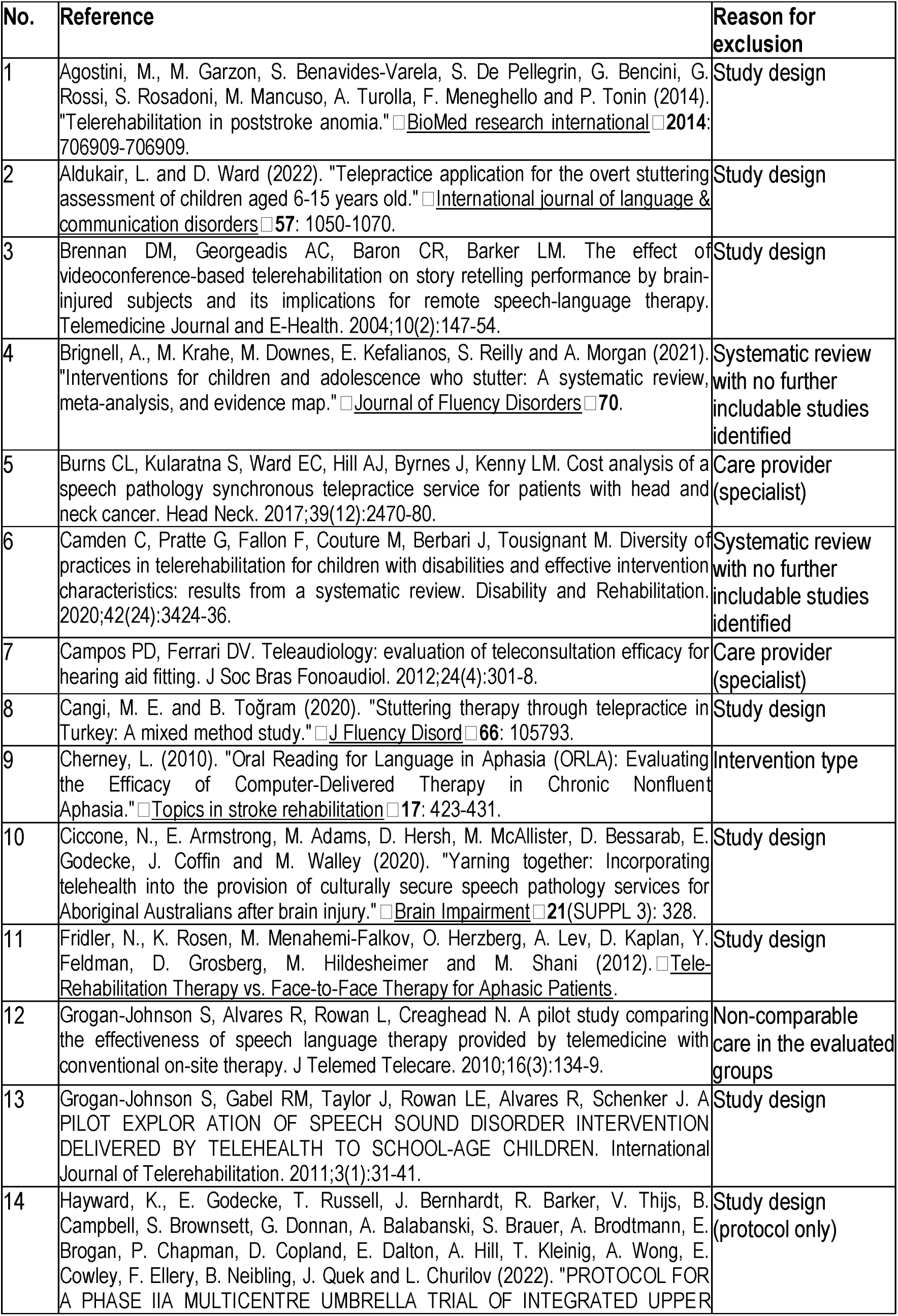
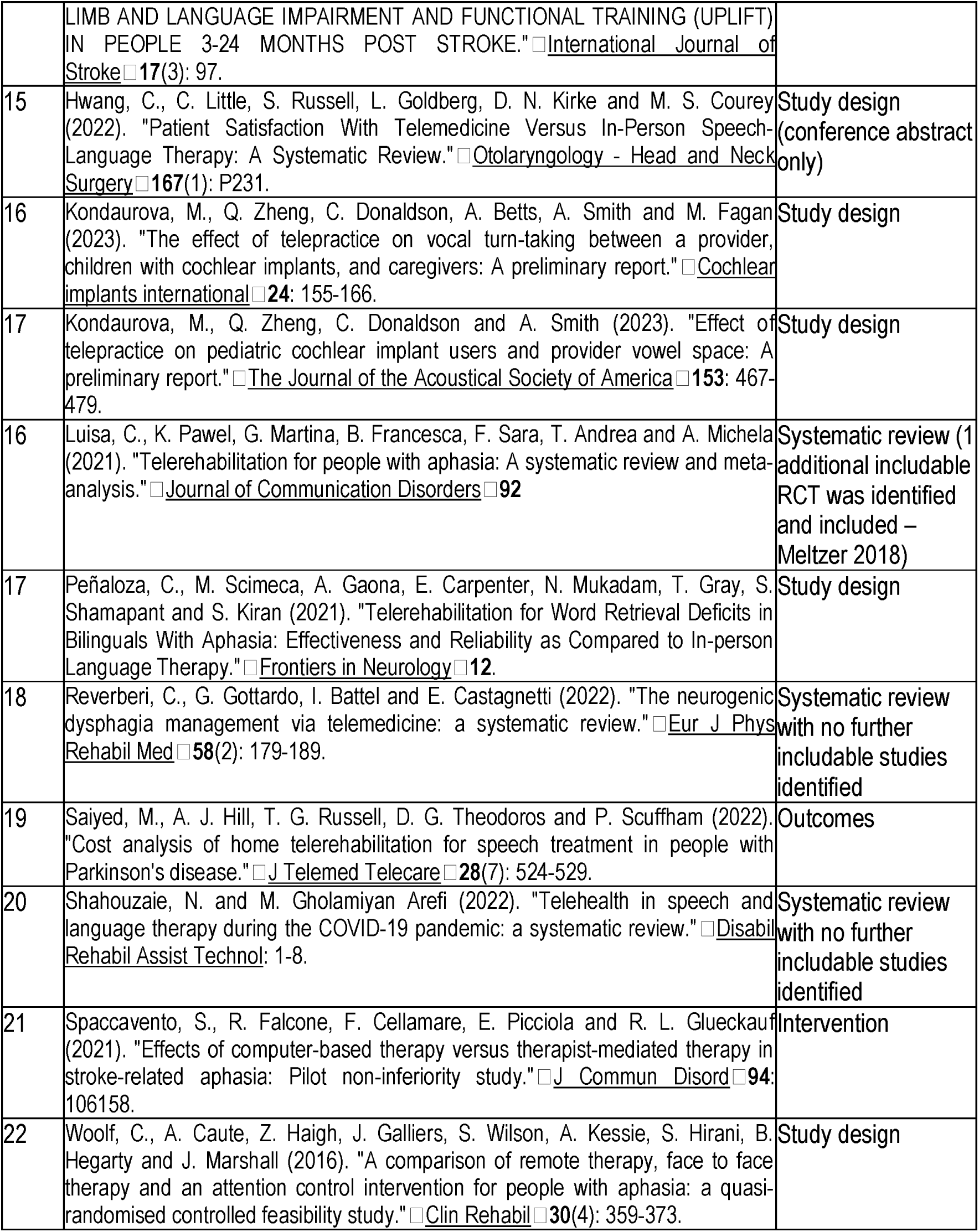

| No. | Reference | Reason for exclusion |
| --- | --- | --- |
| 1 | Agostini, M., M. Garzon, S. Benavides-Varela, S. De Pellegrin, G. Bencini, G. Rossi, S. Rosadoni, M. Mancuso, A. Turolla, F. Meneghello and P. Tonin (2014). "Telerehabilitation in poststroke anomia." <u>BioMed research international</u> **2014**: 706909-706909. | Study design |
| 2 | Aldukair, L. and D. Ward (2022). "Telepractice application for the overt stuttering assessment of children aged 6-15 years old." <u>International journal of language & communication disorders</u> **57**: 1050-1070. | Study design |
| 3 | Brennan DM, Georgeadis AC, Baron CR, Barker LM. The effect of videoconference-based telerehabilitation on story retelling performance by brain-injured subjects and its implications for remote speech-language therapy. <u>Telemedicine Journal and E-Health</u>. 2004;10(2):147-54. | Study design |
| 4 | Brignell, A., M. Krahe, M. Downes, E. Kefalianos, S. Reilly and A. Morgan (2021). "Interventions for children and adolescence who stutter: A systematic review, meta-analysis, and evidence map." <u>Journal of Fluency Disorders</u> **70**. | Systematic review with no further includable studies identified |
| 5 | Burns CL, Kularatna S, Ward EC, Hill AJ, Byrnes J, Kenny LM. Cost analysis of a speech pathology synchronous telepractice service for patients with head and neck cancer. <u>Head Neck</u>. 2017;39(12):2470-80. | Care provider (specialist) |
| 6 | Camden C, Pratte G, Fallon F, Couture M, Berbari J, Tousignant M. Diversity of practices in telerehabilitation for children with disabilities and effective intervention characteristics: results from a systematic review. <u>Disability and Rehabilitation</u>. 2020;42(24):3424-36. | Systematic review with no further includable studies identified |
| 7 | Campos PD, Ferrari DV. Teleaudiology: evaluation of teleconsultation efficacy for hearing aid fitting. <u>J Soc Bras Fonoaudiol</u>. 2012;24(4):301-8. | Care provider (specialist) |
| 8 | Cangi, M. E. and B. Toğram (2020). "Stuttering therapy through telepractice in Turkey: A mixed method study." <u>J Fluency Disord</u> **66**: 105793. | Study design |
| 9 | Cherney, L. (2010). "Oral Reading for Language in Aphasia (ORLA): Evaluating the Efficacy of Computer-Delivered Therapy in Chronic Nonfluent Aphasia." <u>Topics in stroke rehabilitation</u> **17**: 423-431. | Intervention type |
| 10 | Ciccone, N., E. Armstrong, M. Adams, D. Hersh, M. McAllister, D. Bessarab, E. Godecke, J. Coffin and M. Walley (2020). "Yarning together: Incorporating telehealth into the provision of culturally secure speech pathology services for Aboriginal Australians after brain injury." <u>Brain Impairment</u> **21**(SUPPL 3): 328. | Study design |
| 11 | Fridler, N., K. Rosen, M. Menahemi-Falkov, O. Herzberg, A. Lev, D. Kaplan, Y. Feldman, D. Grosberg, M. Hildesheimer and M. Shani (2012). <u>Tele-Rehabilitation Therapy vs. Face-to-Face Therapy for Aphasic Patients</u>. | Study design |
| 12 | Grogan-Johnson S, Alvares R, Rowan L, Creaghead N. A pilot study comparing the effectiveness of speech language therapy provided by telemedicine with conventional on-site therapy. <u>J Telemed Telecare</u>. 2010;16(3):134-9. | Non-comparable care in the evaluated groups |
| 13 | Grogan-Johnson S, Gabel RM, Taylor J, Rowan LE, Alvares R, Schenker J. A PILOT EXPLORATION OF SPEECH SOUND DISORDER INTERVENTION DELIVERED BY TELEHEALTH TO SCHOOL-AGE CHILDREN. <u>International Journal of Telerehabilitation</u>. 2011;3(1):31-41. | Study design |
| 14 | Hayward, K., E. Godecke, T. Russell, J. Bernhardt, R. Barker, V. Thijs, B. Campbell, S. Brownsett, G. Donnan, A. Balabanski, S. Brauer, A. Brodtmann, E. Brogan, P. Chapman, D. Copland, E. Dalton, A. Hill, T. Kleinig, A. Wong, E. Cowley, F. Ellery, B. Neibling, J. Quek and L. Churilov (2022). "PROTOCOL FOR A PHASE IIA MULTICENTRE UMBRELLA TRIAL OF INTEGRATED UPPER | Study design (protocol only) |
|  | LIMB AND LANGUAGE IMPAIRMENT AND FUNCTIONAL TRAINING (UPLIFT) IN PEOPLE 3-24 MONTHS POST STROKE." □ <u>International Journal of Stroke</u> □ **17**(3): 97. |  |
| 15 | Hwang, C., C. Little, S. Russell, L. Goldberg, D. N. Kirke and M. S. Courey (2022). "Patient Satisfaction With Telemedicine Versus In-Person Speech-Language Therapy: A Systematic Review." □ <u>Otolaryngology - Head and Neck Surgery</u> □ **167**(1): P231. | Study design<br>(conference abstract only) |
| 16 | Kondaurova, M., Q. Zheng, C. Donaldson, A. Betts, A. Smith and M. Fagan (2023). "The effect of telepractice on vocal turn-taking between a provider, children with cochlear implants, and caregivers: A preliminary report." □ <u>Cochlear implants international</u> □ **24**: 155-166. | Study design |
| 17 | Kondaurova, M., Q. Zheng, C. Donaldson and A. Smith (2023). "Effect of telepractice on pediatric cochlear implant users and provider vowel space: A preliminary report." □ <u>The Journal of the Acoustical Society of America</u> □ **153**: 467-479. | Study design |
| 16 | Luisa, C., K. Pawel, G. Martina, B. Francesca, F. Sara, T. Andrea and A. Michela (2021). "Telerehabilitation for people with aphasia: A systematic review and meta-analysis." □ <u>Journal of Communication Disorders</u> □ **92** | Systematic review (1 additional includable RCT was identified and included – Meltzer 2018) |
| 17 | Peñaloza, C., M. Scimeca, A. Gaona, E. Carpenter, N. Mukadam, T. Gray, S. Shamapant and S. Kiran (2021). "Telerehabilitation for Word Retrieval Deficits in Bilinguals With Aphasia: Effectiveness and Reliability as Compared to In-person Language Therapy." □ <u>Frontiers in Neurology</u> □ **12**. | Study design |
| 18 | Reverberi, C., G. Gottardo, I. Battel and E. Castagnetti (2022). "The neurogenic dysphagia management via telemedicine: a systematic review." □ <u>Eur J Phys Rehabil Med</u> □ **58**(2): 179-189. | Systematic review<br>with no further includable studies identified |
| 19 | Saiyed, M., A. J. Hill, T. G. Russell, D. G. Theodoros and P. Scuffham (2022). "Cost analysis of home telerehabilitation for speech treatment in people with Parkinson's disease." □ <u>J Telemed Telecare</u> □ **28**(7): 524-529. | Outcomes |
| 20 | Shahouzaie, N. and M. Gholamiyan Arefi (2022). "Telehealth in speech and language therapy during the COVID-19 pandemic: a systematic review." □ <u>Disabil Rehabil Assist Technol</u>: 1-8. | Systematic review<br>with no further includable studies identified |
| 21 | Spaccavento, S., R. Falcone, F. Cellamare, E. Picciola and R. L. Glueckauf (2021). "Effects of computer-based therapy versus therapist-mediated therapy in stroke-related aphasia: Pilot non-inferiority study." □ <u>J Commun Disord</u> □ **94**: 106158. | Intervention |
| 22 | Woolf, C., A. Caute, Z. Haigh, J. Galliers, S. Wilson, A. Kessie, S. Hirani, B. Hegarty and J. Marshall (2016). "A comparison of remote therapy, face to face therapy and an attention control intervention for people with aphasia: a quasi-randomised controlled feasibility study." □ <u>Clin Rehabil</u> □ **30**(4): 359-373. | Study design |

## Appendix 3 – PRISMA 2020 Checklist

| Section and Topic | Item # | Checklist item | Location where item is reported |
| --- | --- | --- | --- |
| **TITLE** |  |  |  |
| Title | 1 | Identify the report as a systematic review. | Title |
| **ABSTRACT** |  |  |  |
| Abstract | 2 | See the PRISMA 2020 for Abstracts checklist. | Abstract |
| **INTRODUCTION** |  |  |  |
| Rationale | 3 | Describe the rationale for the review in the context of existing knowledge. | Introduction section. |
| Objectives | 4 | Provide an explicit statement of the objective(s) or question(s) the review addresses. | Introduction section. |
| **METHODS** |  |  |  |
| Eligibility criteria | 5 | Specify the inclusion and exclusion criteria for the review and how studies were grouped for the syntheses. | Methods: inclusion criteria section. |
| Information sources | 6 | Specify all databases, registers, websites, organisations, reference lists and other sources searched or consulted to identify studies. Specify the date when each source was last searched or consulted. | Methods: search strategies section + Appendix (search strings) |
| Search strategy | 7 | Present the full search strategies for all databases, registers and websites, including any filters and limits used. | Appendix 1: Searches |
| Selection process | 8 | Specify the methods used to decide whether a study met the inclusion criteria of the review, including how many reviewers screened each record and each report retrieved, whether they worked independently, and if applicable, details of automation tools used in the process. | Methods: study selection and screening section. |
| Data collection process | 9 | Specify the methods used to collect data from reports, including how many reviewers collected data from each report, whether they worked independently, any processes for obtaining or confirming data from study investigators, and if applicable, details of automation tools used in the process. | Methods: data extraction section. |
| Data items | 10a | List and define all outcomes for which data were sought. Specify whether all results that were compatible with each outcome domain in each study were sought (e.g. for all measures, time points, analyses), and if not, the methods used to decide which results to collect. | Methods: data extraction section. |
|  | 10b | List and define all other variables for which data were sought (e.g. participant and intervention characteristics, funding sources). Describe any assumptions made about any missing or unclear information. | Methods: data extraction section. |
| Study risk of bias assessment | 11 | Specify the methods used to assess risk of bias in the included studies, including details of the tool(s) used, how many reviewers assessed each study and whether they worked independently, and if applicable, details of automation tools used in the process. | Methods: Assessment of the risk of bias section. |
| Effect measures | 12 | Specify for each outcome the effect measure(s) (e.g. risk ratio, mean difference) used in the synthesis or presentation of results. | Methods: Measurement of effect and data synthesis section |
| Synthesis methods | 13a | Describe the processes used to decide which studies were eligible for each synthesis (e.g. tabulating the study intervention characteristics and comparing against the planned groups for each synthesis (item #5)). | Methods: Measurement of effect and data synthesis section |
|  | 13b | Describe any methods required to prepare the data for presentation or synthesis, such as handling of missing summary statistics, or data conversions. | Methods: Measurement of effect and data synthesis section |
|  | 13c | Describe any methods used to tabulate or visually display results of individual studies and syntheses. | Methods: Measurement of effect and data synthesis section |
|  | 13d | Describe any methods used to synthesize results and | Methods: Measurement of |
|  |  | provide a rationale for the choice(s). If meta-analysis was performed, describe the model(s), method(s) to identify the presence and extent of statistical heterogeneity, and software package(s) used. | effect and data synthesis section |
|  | 13e | Describe any methods used to explore possible causes of heterogeneity among study results (e.g. subgroup analysis, meta-regression). | Methods: Measurement of effect and data synthesis section |
|  | 13f | Describe any sensitivity analyses conducted to assess robustness of the synthesized results. | Methods: Measurement of effect and data synthesis section |
| Reporting bias assessment | 14 | Describe any methods used to assess risk of bias due to missing results in a synthesis (arising from reporting biases). | Not applicable. |
| Certainty assessment | 15 | Describe any methods used to assess certainty (or confidence) in the body of evidence for an outcome. | Not applicable. |
| **RESULTS** |  |  |  |
| Study selection | 16a | Describe the results of the search and selection process, from the number of records identified in the search to the number of studies included in the review, ideally using a flow diagram. | Results: search results section |
|  | 16b | Cite studies that might appear to meet the inclusion criteria, but which were excluded, and explain why they were excluded. | Appendix B |
| Study characteristics | 17 | Cite each included study and present its characteristics. | Results: summary of included trials |
| Risk of bias in studies | 18 | Present assessments of risk of bias for each included study. | Results: risk of bias section |
| Results of individual studies | 19 | For all outcomes, present, for each study: (a) summary statistics for each group (where appropriate) and (b) an effect estimate and its precision (e.g. confidence/credible interval), ideally using structured tables or plots. | Results: TH vs F2F SLP for patients with stuttering section<br>Results: TH vs F2F SLP for patients with Parkinson's disease section |
| Results of syntheses | 20a | For each synthesis, briefly summarise the characteristics and risk of bias among contributing studies. | Results: TH vs F2F SLP for patients with stuttering section<br>Results: TH vs F2F SLP for patients with Parkinson's disease section |
|  | 20b | Present results of all statistical syntheses conducted. If meta-analysis was done, present for each the summary estimate and its precision (e.g. confidence/credible interval) and measures of statistical heterogeneity. If comparing groups, describe the direction of the effect. | Results: TH vs F2F SLP for patients with stuttering section<br>Results: TH vs F2F SLP for patients with Parkinson's disease section |
|  | 20c | Present results of all investigations of possible causes of heterogeneity among study results. | Results: TH vs F2F SLP for patients with stuttering section<br>Results: TH vs F2F SLP for patients with Parkinson's disease section |
|  | 20d | Present results of all sensitivity analyses conducted to assess the robustness of the synthesized results. | No sensitivity analyses conducted. |
| Reporting biases | 21 | Present assessments of risk of bias due to missing results (arising from reporting biases) for each synthesis assessed. | Not applicable |
| Certainty of evidence | 22 | Present assessments of certainty (or confidence) in the body of evidence for each outcome assessed. | Not applicable |
| **DISCUSSION** |  |  |  |
| Discussion | 23a | Provide a general interpretation of the results in the context of other evidence. | Discussion section |
|  | 23b | Discuss any limitations of the evidence included in the review. | Discussion section |
|  | 23c | Discuss any limitations of the review processes used. | Discussion section |
|  | 23d | Discuss implications of the results for practice, policy, and future research. | Discussion section |
| **OTHER INFORMATION** |  |  |  |
| Registration and protocol | 24a | Provide registration information for the review, including register name and registration number, or state that the review was not registered. | Protocol for the overall review was developed a priori but not registered. |
|  | 24b | Indicate where the review protocol can be accessed, or state that a protocol was not prepared. | From study authors. |
|  | 24c | Describe and explain any amendments to information provided at registration or in the protocol. | Reported in the relevant methods section. |
| Support | 25 | Describe sources of financial or non-financial support for the review, and the role of the funders or sponsors in the review. | Funding and COI statement (Appendix) |
| Competing interests | 26 | Declare any competing interests of review authors. | Funding and COI statement (Appendix) |
| Availability of data, code and other materials | 27 | Report which of the following are publicly available and where they can be found: template data collection forms; data extracted from included studies; data used for all analyses; analytic code; any other materials used in the review. | From study authors. |

## Appendix 4 – Funding and COI statements

### COI and funding disclosures

This systematic review was commissioned by the Department of Health and Aged Care, Canberra, Australia, as part of a series of systematic reviews on the effectiveness of telehealth within primary care in 2020-21 and their update in 2023. The funder was involved in establishing the parameters of the study question (PICO). The funder was not involved in the conduct, analysis, or interpretation of the systematic review, or in the decision to submit the manuscript for publication.

The authors report no other actual or potential conflicts of interest.

